# Evaluation of COVID 19 infection in 279 cancer patients treated during a 90-day period in 2020 pandemic

**DOI:** 10.1101/2020.05.26.20102889

**Authors:** Mozaffar Aznab

**Affiliations:** Kermanshsh University of Medical science

**Keywords:** COVID-19, Cancer, Treatment, Solid Tumor, Hematology Malignancy

## Abstract

**Background:** The aim of this study was investigation of COVID-19 disease and its outcome in cancer patients who needed treatment, in a 90-day period.

**Methods:** Cancer patient who required treatment, were evaluated for potential COVID-19 infection in a 90-day period, starting from beginning of this epidemic in Iran, January, to April 19, 2020. For treatment of solid tumor patients, if they did not have symptoms related to COVID-19, just chest X-ray was requested. If they showed COVID-19 related symptoms, High Resolution CT scan of lungs was requested. For hematology cancer patients, PCR test for COVID-19 infection was requested as well. Protection measures were considered for personnel of oncology wards.

**Results:** In this study, 279 patients were followed up in this 90-day period. No COVID-19 infection was observed in 92 cases of breast cancer, 72 cases of colon cancer, 14 cases of gastric cancer and 12 cases of pancreaticobiliary cancer.However, in 11 cases of lung cancer, 5 cases brain tumors and 12 cases ovarian cancer; 3 case of COVID-19 were observed. In the hematology cancers group, which included 14 cases of Hodgkin Lymphoma, 23 cases of lymphoproliferative disorder, 12 cases of acute leukemia and 12 cases of multiple myeloma; three of COVID-19 were observed.

**Conclusion:** Patients with cancer who need treatment can be treated by taking some measures. These measures include observing individual and collective protection principles in patients and health-care personnel, increasing patients awareness particularly about self-care behavior, performing a COVID-19 test, and taking a chest X ray, before the treatment starts

## Introduction

Unfortunately, since December 2019 the world has faced a new Coronavirus epidemic which has been started from Wuhan, China. In 2020 the World Health Organization (WHO) considered this infection a serious global health problem and announced it as a pandemic due to its outbreak all around the world. The most serious complication caused by this infection was a severe acute respiratory syndrome which led to lots of deaths [1]. The most important symptoms presenting this infection were cough, shortness of breath and sore throat; after a while, other symptoms including diarrhea, vomiting, changes in taste and smell, and headache were reported too [2]. The most important complication which results in death is severe acute respiratory syndrome. The number of resulted deaths in April 2020 was more than 45000 [3]. The incubation period of COVID-19 infection is reported to be 2 to 14 days; 5.2 days on average since transmission time [4, 5]. In some studies, the average incubation time is reported 6.4 days. Diagnosis of this infection is done using Real-time PCR test [6]. According a report from CDC [7], high-risk groups for being infected or showing severe illness of COVID-19 are elderly adults and people of any age who have serious underlying medical conditions, particularly if it is not well controlled. Based on the CDC’s report, those who are at high-risk for severe illness are: people 60 years or older, people who live in nursing homes, people with chronic lung disease or moderate to severe asthma, people who have serious heart conditions, people who are immunocompromised (including patient who undergo cancer treatment, smokers, people who had bone marrow or organ transplantation, and people with prolonged use of corticosteroids). Moreover, people with severe obesity, diabetic patients, people with chronic liver disease and people with chronic kidney are among other high-risk groups. According to some reports, cancer patients are at a higher risk for severe illness from COVID-19, compared to the public [8]. Yet, there is no accurate and concise instruction for cancer patients to deal with this epidemic. COVID-19 is a challenge for oncology and hematology specialists. There are some suggestions about managing these patients, including instructions in ESMO site [9]. According to these suggestions, patients are divided into groups called Priority A, Priority B, and Priority C. These divisions are based on some facts; including if the cancer patient has an active cancer which is life-threatening, if they are at an average risk, or if they do not currently have active cancer. The aim of our study was to investigate COVID-19 illness and its outcome in cancer patients who needed treatment.

## Methods and Material

Cancer patients who required treatment, were evaluated for potential COVID-19 infection in a 90-day period, starting from beginning of this epidemic in Iran, January 21, 2020 to April 19, 2020. The treatments were injection therapies including chemotherapy, Bevacizumab, Rituximab, Trastuzumab, Rituximab, Sunitinib, Sorafenib, Everolimus, Erlotinib, CCNU *(Lomustine)*, Procarbazin and etc. Which were used based on the cancer’s type. Patients divided in to two groups of solid tumors and hematology cancers. In solid tumor patients, if they did not have any symptoms related to COVID-19 or other infection symptoms, just chest X-ray was requested before starting treatment. If they showed COVID-19 related symptoms, High Resolution CT scan of lungs was requested. Otherwise the cancer treatment would be started. In hematology cancers group, in addition to above mentioned tests, real-time PCR test for COVID-19 was requested before starting the treatment. Unfortunately, performing PCR test was not always possible. All patients and their families were added to a telegram group to receive educations related to protection and self-care, since beginning of this epidemic. In this group they were trained about protective measures such as using disinfectants, masks, use soap and water to wash hands, and other hygiene methods. Since the very beginning of the epidemic we suggested patients and their families to use masks, we suggested it was necessary for everyone to use masks, and we considered protection measures for personnel of oncology and clinic wards [10]. We paid special attention to increase peoples’ awareness particularly about self-care behaviors. These people included patients, their families and health-care personnel. Chemotherapy treatments were performed as usual. No clinical trials or new drugs have been used during this time. For patients, findings related to cancer and QVID-19 infection were described, and if treatment was agreed upon, treatment consent was obtained from the patients.

## Results

After this 90-day period, 279 patients with cancer who were undergoing cancer treatments were evaluated. 218 patients had solid tumors and 61 patients had hematological cancers. Among these 218 solid tumor patients, 92 cases had breast cancer whose treatment was either started before the epidemic and continued throughout this period, or the treatment was started during the epidemic. The staging of patients was as follows; 5 patients in stage I, 29 patients in stage II, 37 patients in stage III, and 21 patients in metastatic stage (some of these patients, referred to as metastases, were patients with recurrence of the disease). Twenty-one of the 29 patients were receiving Herceptin during the epidemic, and the usual chemotherapy was completed before the epidemic began. For breast cancer patients, treatment was started with the same usual dose and at the same usual intervals of every 3 weeks. We had no COVID-19 positive case in these patients. Among 72 patients with colon cancer, 11 patients were in stage II, 36 patients were in stage III, and 25 patients were in metastatic stage (some of these cases were relapsed patients). For these patients, treatment was either started during the epidemic or continued during the epidemic. Treatment was done once every two weeks (FOLFOX4, FOLFIRI) and in metastastic cases Bevacizumab (Stivant) or Cetuximab was used. In majority of stage II patients, a three-week regimen of (Capacitabin/Oxalliplatin) was used. We had no COVID-19 positive case in these patients (table1). There were 11 patients with lung cancer; of which 7 patients were taking chemotherapy, and 3 patients were taking Erlotinib. Among these 11 cases of lung cancer, one case of COVID-19 was observed. The patient developed a COVID-19 infection before cancer treatment starts, and died as a result. In 12 patients with pancreaticobiliary cancer no COVID-19 positive case was reported. We had 5 patients with glioblastoma multiforme; in this group we had one case of COVID-19 infection. The patient was under the ventilator device and later died as a result of COVID-19 infection. In 12 cases with ovarian cancer, one suspected case with positive lung CT scan was observed, but COVID-19 PCR test for this patient was negative Characteristic of patients with other cancers are listed in table 2. In hematology cancer group we had 14 cases of Hodgkin’s disease who were receiving treatments; 2 patients were in stage I, 4 patients in stage II and 4 patients in stage III. 2 cases were relapsed patients, one was refractory, and one patient was in stage IV. The treatment regimen was ABVD (Adriamycin,Bleomycin,Vinblastin,Dacarbazin) every two weeks. No COVID-19 case was reported in this group. We had 23 cases of lymphoproliferative disorder of which 7 cases received CHOP-Rituximab (Cyclophosphamide,Vincristine,Adriamycin,Prednisolon-Rituximab) treatment, 5 cases received Rituximab, one case received CVP-Rituximab(Cyclophosphamide,Vincristine,Prednisolon-Ritoximab), 3 cases Hyper C-VAD, 4 cases Fludarabin/Cyclophosmade/Retuximab and 2 cases received Bendamustine /Prednisolone (table 3). There was one COVID-19 positive case with pulmonary clinical symptoms in a patient with cerebral lymphoma which now is receiving supportive treatments. There were 12 cases of acute leukemia, 5 of which were lymphoblastic leukemia. Among these four cases, one died due to lack of response to the treatment; one patient has been diagnosed with COVID-19 during neutropenia phase and died. Three other patients have no specific problems. There were 7 cases of myeloblastic leukaemia, 2 of which had a history of CML and using Nilotinib; which entered the blastic phase and were receiving (7+3 days) Cytosine Arabinoside / Daunorubicin Currently, these two patients are in good conditions. There were two cases of de novo acute myeloid leukemia. One of them was relapsed before the epidemic began, it was resistant to therapy, and the patient died due to leukostasis. There were three cases of M3 Acute Promyelocytic leukaemia which are receiving Arsenic and do not have any particular problems [11]. We had 12 cases of myeloma in this 90-day period. Three of them received Melphalan and Prednisolone. Among them five cases received proteasome inhibitor (Alvocade); in this group there was one COVID-19 positive case which is in good general condition and has no symptoms. There were 4 cases of myeloma which underwent C-VAD regimen (Cyclophosphamide-vincristine adriamycin dexamethasone) [12]. One of them suddenly developed fever and respiratory distress and died within 24 hours before the treatment begins. The cause of death is not clear and it was not possible to be evaluated by COVID-19 PCR test.

**Table 1;.**
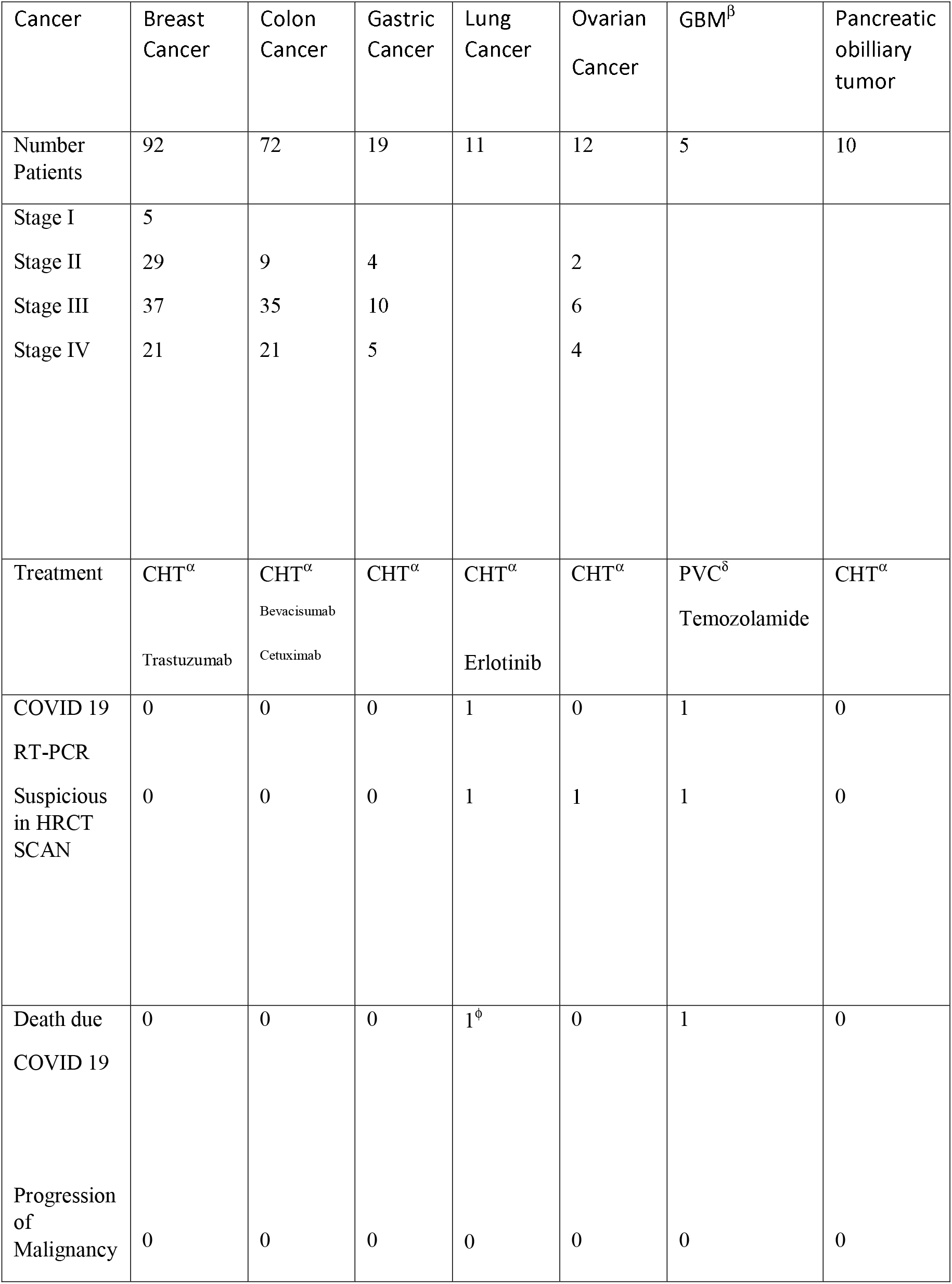
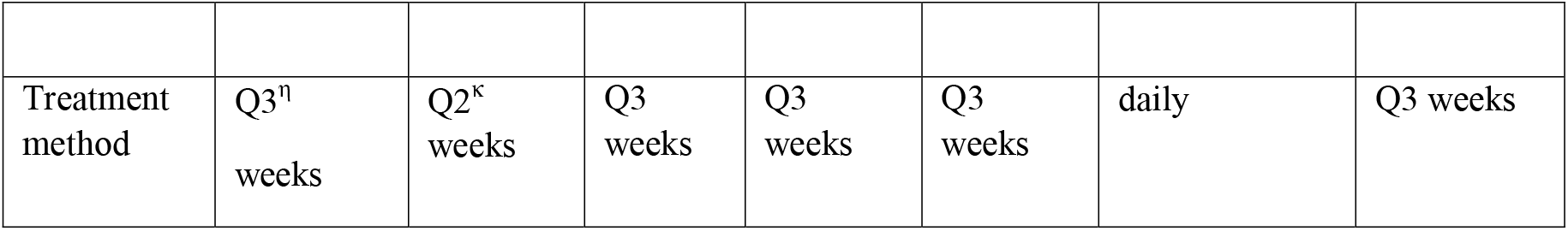
the relationship between cancer treatment and the prevalence of COVID 19 ϕ:No treatment for malignancy α;CHT: Chemotherapy β;GBM: Glioblastoma Multiform δ: Procarbazine, Lomustine(CCNU), and Vincristine (PCV) κ;Q 2 weeks:Once every 2 weeks η; Q3 weeks::Once every 3 weeks

**Table 2;.**
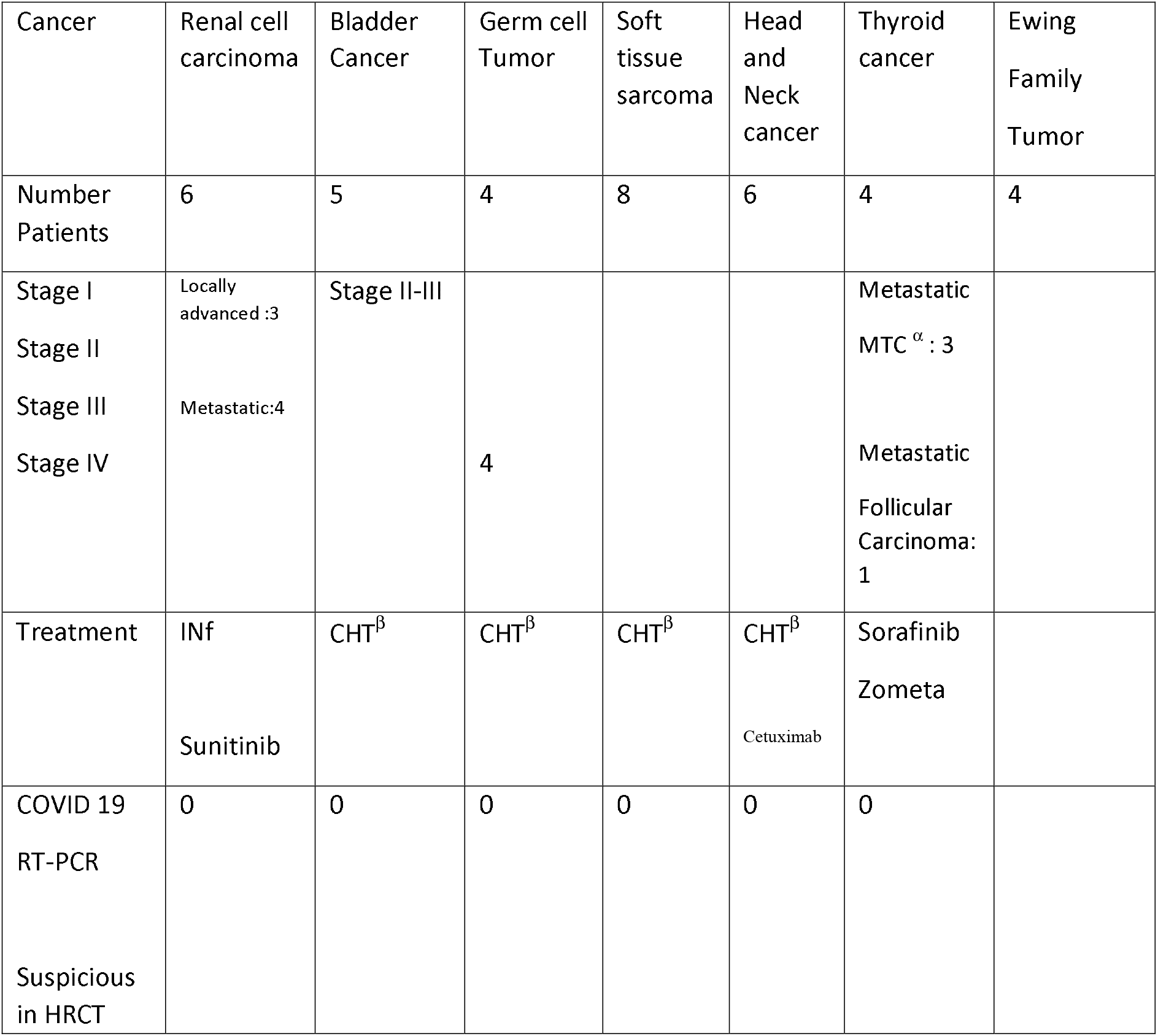
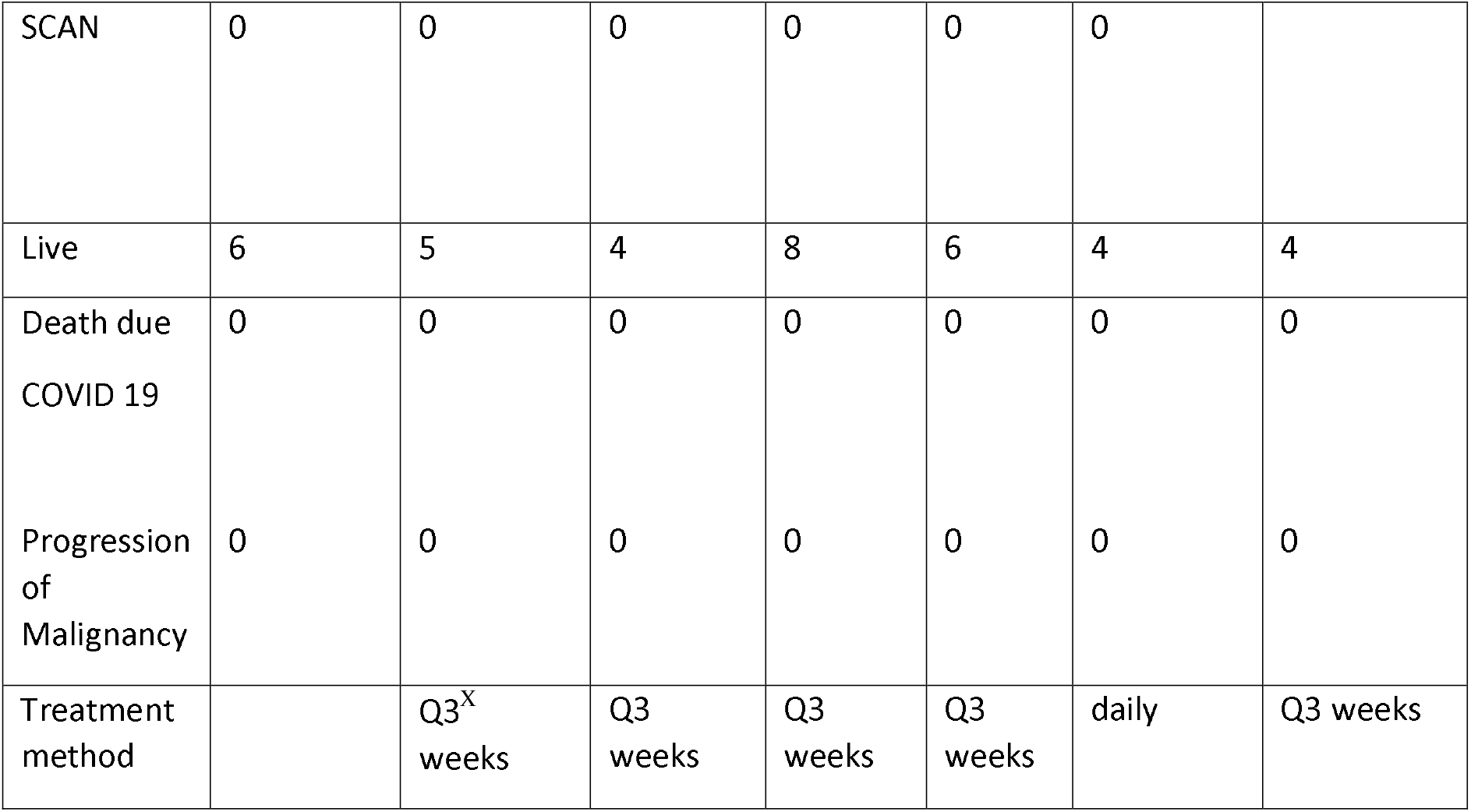
the relationship between other cancer treatment and the prevalence of COVID 19 α^;^MTC: Medullary Thyroid Carcinoma B;CHT: Chemotherapy X;Q3 weeks: Once every three week

**Table 3;.**
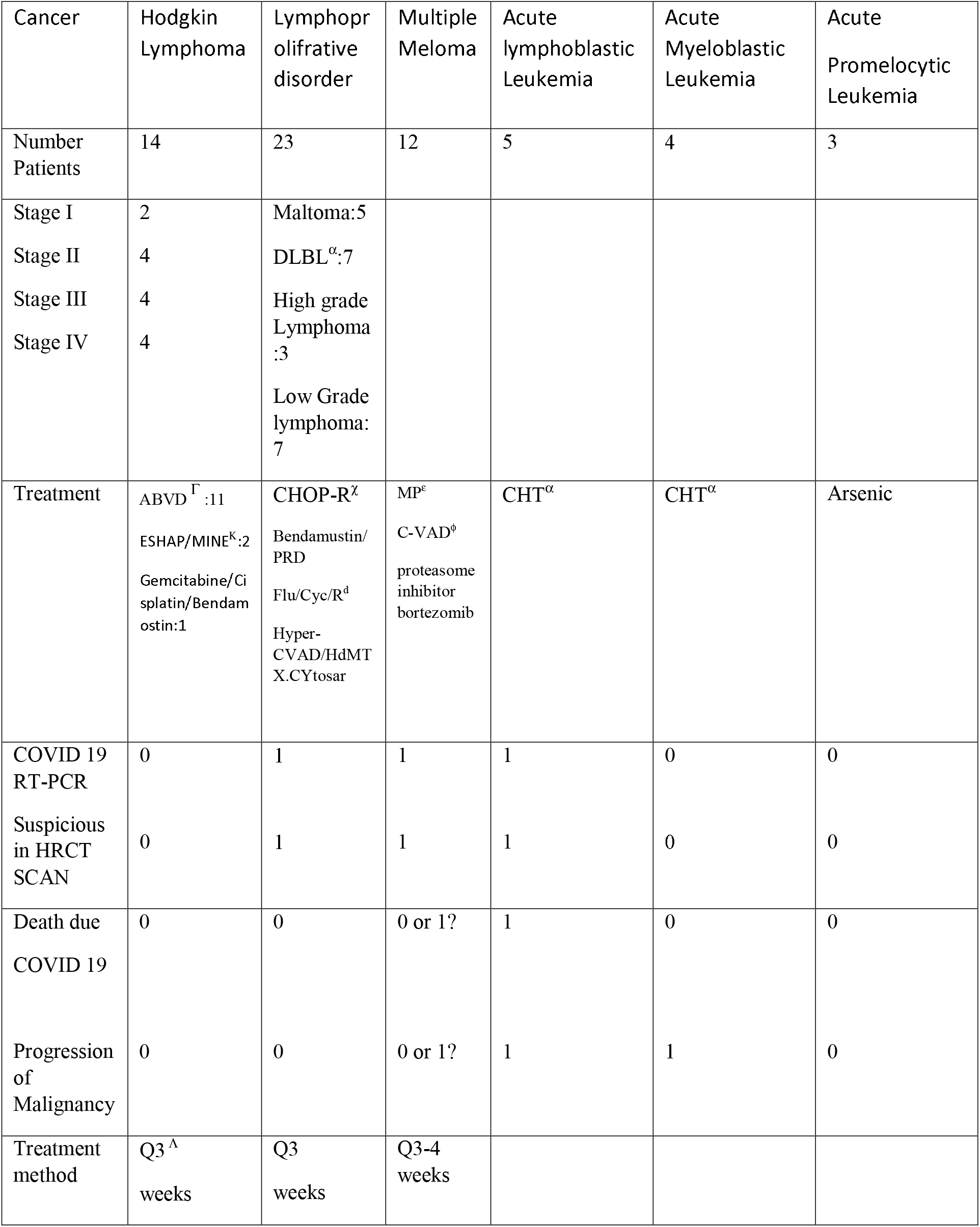
the relationship between hematology malignancy treatment and the prevalence of COVID 19 α:Diffuse Large B cell Lymphoma χ:Cyclophosphamied, Vincristine, Adriamicine, Prednisolon-Rituximab d: Fludarabin/cyclophosphamide/Rituximab ε:Melphalan prednisolone ϕ:Cyclophosphamide-Vincristine..Dexamethasone..Adriamyin Γ; ABVD:, Adriamicine. Bleomycin. Vinblastin Dacarbazin α CHT:Chemotherapy K;MINE (mesna, ifosfamide, mitoxantrone, and etoposide) alternated with ESHAP (etoposide, methylprednisolone, high-dose cytarabine, and cisplatin Λ; Q 3-4 week: Once every 3-4 weeks 1?: The cause of death of a patient in this group was unknown and it was not possible to investigate Quaid 19

## Discussion

Among 11 cases of lung cancer, there was a stage IV lung cancer patient who was COVID-19 positive (in terms of radiology and CT scan).The patient developed a coronavirus infection before the cancer treatment and died. We had 5 patients with glioblastoma multiforme, in this group we had one case of COVID-19 infection that was under the ventilator device resulted death was observed. In 12 cases with ovarian cancer, there was one patient with ovarian cancer which was a suspected case in terms CT scan. Following this patient, the general condition is good, the test results of RT-PCR of COVID-19 negative was reported and in the new CT scan of the lungs, there was no specific sign in favor of COVID-19 infection and the next course of treatment was started. In hematology cancers group, we had a patient with refractory acute lymphoblastic leukemia who was positive for COVID-19 and died as its result. Among the 5 of the 12 cases of Multiple Myeloma that received proteasome inhibitor (Alvocade); there was one COVID-19 positive case which currently in good general condition and has no symptoms. One of the patients in the Multiple Myeloma group who received (C-CVD, Thalidomide) chemotherapy regimen suddenly developed a fever and respiratory distress and died within 24 hours before starting the second course of treatment, while on the day of admission, the general condition was good. The cause of death is not clear and it was not possible to be evaluated by COVID-19 PCR test. We had a patient with cerebral lymphoma, which was a suspected case in terms CT scan, but it was negative for COVID-19 PCR test, that’s right now, the general condition of the patient is good and there is no particular problem in the field of infection of COVID 19 and the candidate is continuing treatment. The characteristic of patients with COVID-19 are shown in Table 4.

**Table 4;.**
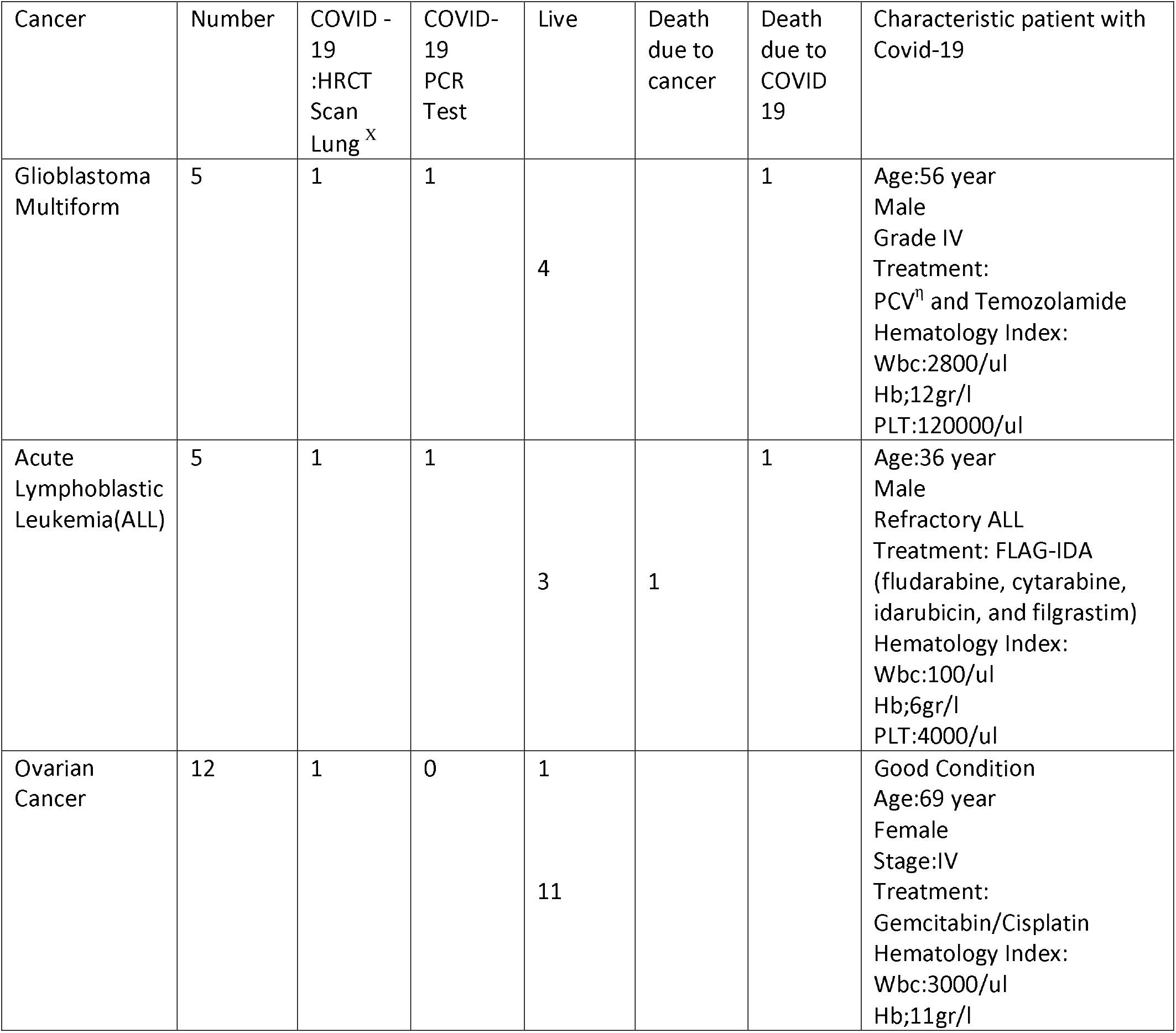
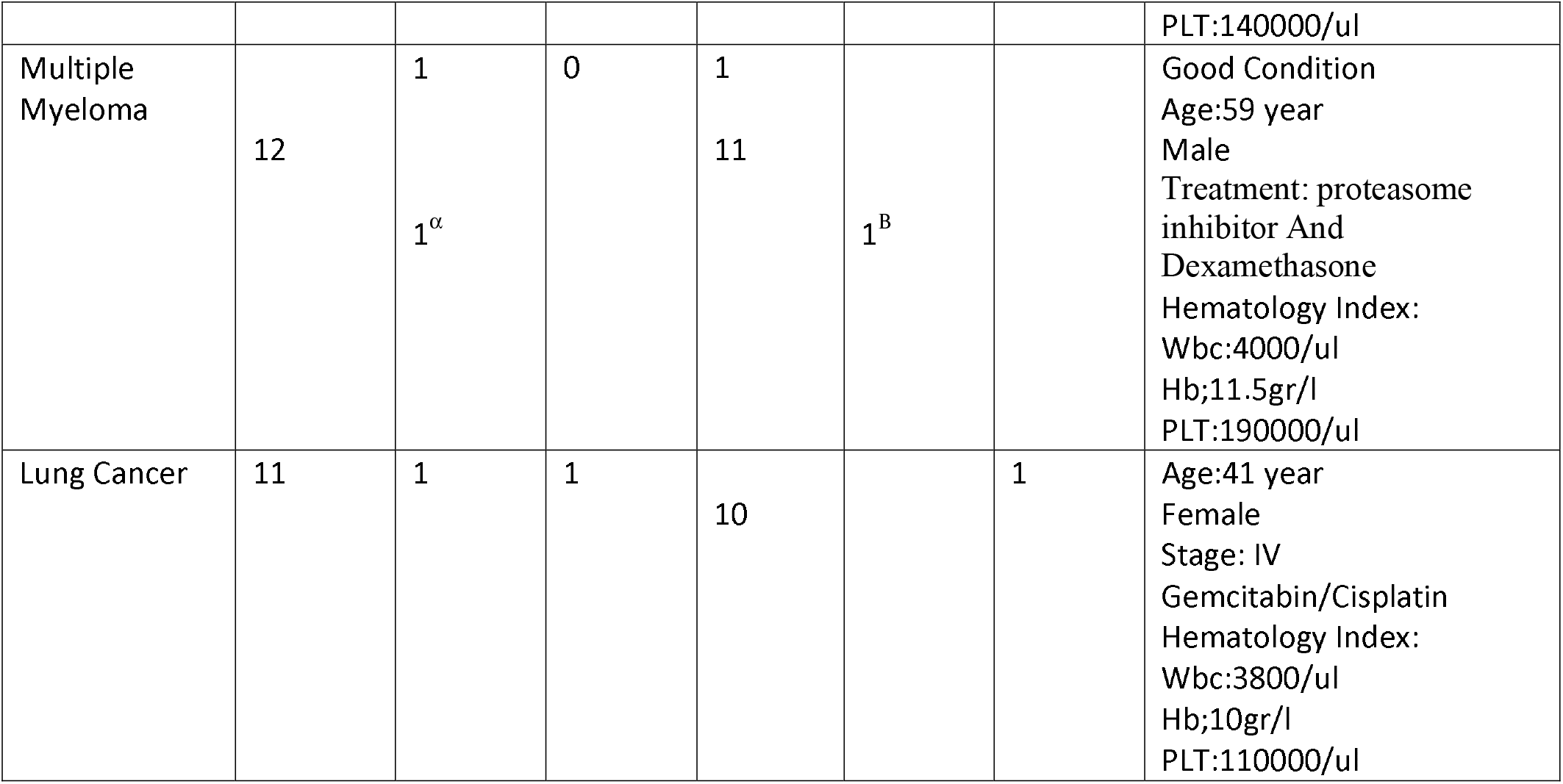
Cancer patients with infection COVID-19 α:Suspicious B; The cause of death of a patient in this group was unknown and it was not possible to investigate QVID 19 X: High resolution CT scan Lung η:Procarbazine, Lomustine(CCNU), and Vincristine (PCV)

In the solid tumors group, there was one patient with ovarian cancer which was suspected to COVID-19 infection in terms of CT scan, but it was negative for COVID-19 PCR test; she is undergoing follow up and supportive measures. There was an advanced stage IV lung cancer patient, who was COVID-19 positive (in terms of radiology and CT scan), and developed a coronavirus infection before starting the cancer treatment and died. We had a patient with cerebral lymphoma, which was a suspected case in terms CT scan, but it was negative for COVID-19 PCR test. This patient is intubated and under the ventilator device. In 72 cases with colon cancer, there was one COVID-19 positive case, in terms of RT-PCR and Lung High Resolution CT Scan, which now general condition and pulse oximetry of the patient is good. In hematology cancers group, we had a patient with refractory acute lymphoblastic leukemia who was positive for COVID-19 and died as its result. Besides, there was a myeloma patient who died with pulmonary symptoms before the treatment began. Considering the results of this study, we realized that COVID-19 complications were more significant in hematology cancer patients, regarding number of patients and number of treatment sessions. There were fewer complications in solid tumor patients. Therefore, according to our findings in this 90-day period, if all protective and hygienic measures are observed, it is possible for solid tumor patients to continue treatments as usual. However, it is still important to start treatment just for patients who are able to do self-care principals. Our routine for breast cancer was every three weeks. In hematology cancer patients, in lymphoproliferative disorder class, both chemotherapy and Rituximab was done every three weeks and without any changes in dose of treatment. There was no specific problem in patients who used Tyrosine Kinase Inhibitors. The most important complications were observed in acute leukaemia and myeloma; which in this regard, more special and appropriate attention is essential. By visiting patients every day, in week days and holidays, and considering history and type of cancer, and patient profile while visiting, we realized that most of people who are not able to do self-care behavior are likely to have some COVID-19 related problems and may be at a high risk of COVID-19 infection. Another thing which caught our attention during this 90-day period was stress and anxiety [13], which was observed in more than 20% of patients and their families as their only supporters. The same applies for these patients’ physician and the author of this article in terms of stress and anxiety, because the stress was unprecedented in the past 15 year. It is emphasized again, if patient is not able to do self-care behaviors, great deal of care must be taken in treating them.

## Conclusion

COVID-19 is a highly dangerous and stressful disease. However, in COVID-19 pandemic, patients with cancer who need treatment can be treated by taking some necessary measures. By observing protection health principles, promoting self-care behavior, and providing proper equipment and diagnostic tests for patients, their companions and health-care personnel, cancer treatment can be continued as usual and without anxiety. It’s important two consider two issues; preventing neutropenia in patients, and psychiatric counseling for patients and health-care personnel. Presence of a psychiatrist is essential for patients and health-care personnel.

## Data Availability

All patient-related information is provided in this article

---

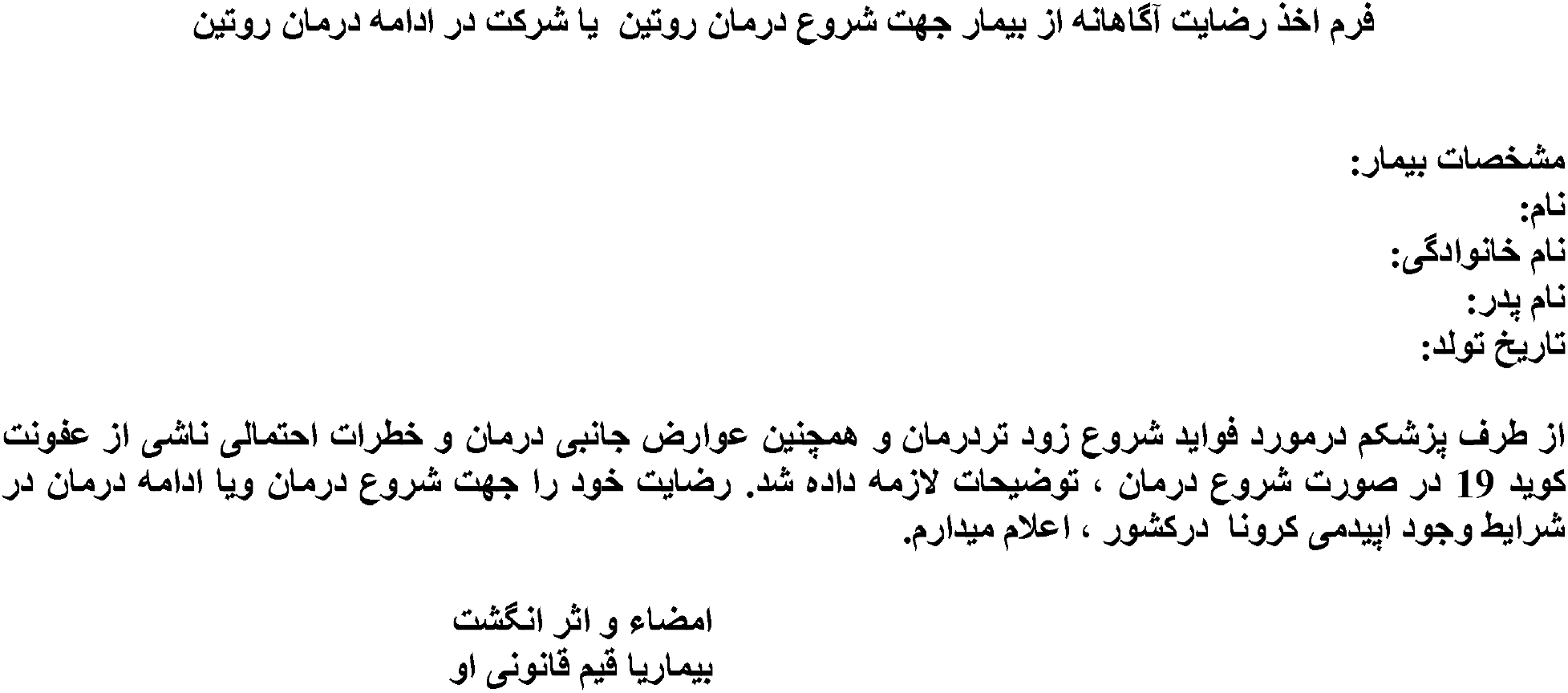

Conscious form of patient consent to initiate routine treatment or to continue routine treatment

Name:

Last Name:

Age:

My doctor explained the benefits of starting early treatment, as well as the side effects of treatment and the potential risks of Quid 19 infection if treatment is started. I express my satisfaction with the start of treatment or the continuation of treatment in the presence of Corona epidemic in the country.

Signature and fingerprint

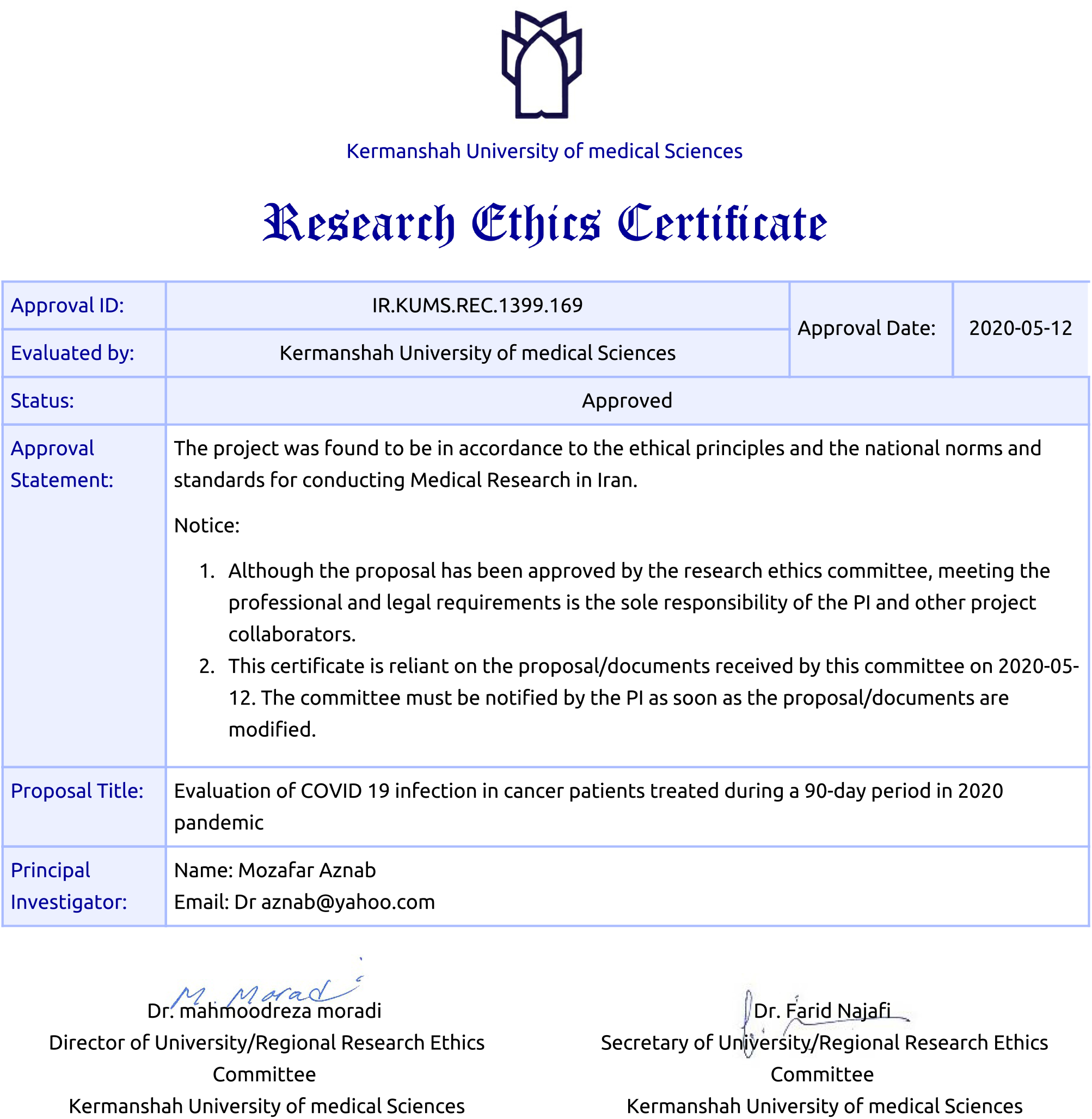

## Reference

1- Zhu N., Zhang D., Wang W. and et al. A novel coronavirus from patients with pneumonia in China, 2019. N Engl J Med. 2020;382:727–733

2- Wu F., Zhao S., Yu B., Chen Y.M., Wang W., Song Z.G. A new coronavirus associated with human respiratory disease in China. Nature. 2020;579:265–269.

3- Johns Hopkins University. Coronavirus COVID-19 Global Cases by the Center for Systems Science and Engineering (CSSE) at Johns Hopkins University. Accessed March 29, 2020. Availableat:https://www.arcgis.com/apps/opsdashboard/index.html#/bda7594740fd40299423467b48e9ecf6

4- Li Q., Guan X., Wu P., Wang X., Zhou L., Tong Y. Early transmission dynamics in Wuhan, China, of novel Coronavirus-infected pneumonia. N Engl J Med. 2020;382:1199–1207.

5- Backer J.A., Klinkenberg D., Wallinga J. Incubation period of 2019 novel coronavirus (2019-nCoV) infections among travellers from Wuhan, China, 20-28 January 2020. Euro Surveill. 2020;25

6- To K.K., Tsang O.T., Chik-Yan Yip C., Chan K.H., Wu T.C., Chan J.M.C. Consistent detection of 2019 novel coronavirus in saliva. Clin Infect Dis. 2020

7- Centers for Disease Control and Prevention. People Who Are at Higher Risk for Severe Illness. Coronavirus Disease 2019 (COVID-19).15 April 2020

8- Liang W, Guan W, Chen R and et al. Cancer patients in SARS-CoV-2 infection: a nationwide analysis in China. Lancet Oncol. 2020 Mar; 21(3):335–337. doi: 10.1016/S1470-2045(20)30096-6

9- Matti Aapro, Alfredo Addeo, Paolo Acierto and et al. Cancer Patient Management During COVID-19 Pandemic. ESMO Recommendations. 17 Apr 2020

10- Cinar P, Kubal T, Freifeld A and et al. Safety at the Time of the COVID-19 Pandemic: How to Keep our Oncology Patients and Healthcare Workers Safe. J Natl Compr Canc Netw. 2020 Apr 15:1–6. doi: 10.6004/jnccn.2020.7572.

11- Mozaffar Aznab, Mansour Rezaei. Induction, consolidation, and maintenance therapies with arsenic as a single agent for acute promyelocytic leukaemia in a 11-year follow-up. Hematol Oncol 2017; 35: 113–117

12- Mozaffar Aznab, Mansour Rezaei, Jafar Navabi, Ali Moieni. Evaluation of low-dose thalidomide as induction and maintenance therapy in patients with multiple myeloma not eligible for stem cell transplantation. APCO 2017; 13: e138-e143

13- Self-Care and Stress Management During the COVID-19 Crisis: Toolkit for Oncology Health Care Professionals. Accessed March 26, 2020. Available at:https://www.nccn.org/members/committees/bestpractices/files/Distress-Management-Clinician-COVID-19.pdf

